# A method enabling computation of linear rates of change of spatial averages on visual field patterns that have varying test locations over time

**DOI:** 10.1101/2025.04.02.25325127

**Authors:** Andrew Turpin, Allison M McKendrick

## Abstract

**Purpose:** To introduce a method for computing linear rates of change in spatial averages for series of visual fields where the test pattern changes over time.

**Method:** We first describe the new method and then verify its performance using simulation on a series of 50 fields where progression occurs at random locations and the test pattern evolves by adding random locations at each visit. We also tested on a series of stable fields, and two “extreme” fields where either all locations were stable except for the 24-2 locations, or all locations were progressing except for the 24-2 locations. The slope estimates using linear regression on an average of a fixed 24-2 pattern and derived from our new method when test locations are added are compared to the known ground truth in the simulations.

**Results:** For random fields and random test patterns, the error in slope estimation differs by less than 0.003 dB/year (paired t.test, p < 0.001) between linear regression slope on MD using the 24-2 pattern and the slope computed using the new method on series where test patterns change.

**Conclusion:** It is possible to calculate linear rates of change on spatial averages that reflect the true underlying rate of change in a series of visual fields where the test pattern changes over time.

## Introduction

It is current practice for clinical visual field (VF) testing (perimetry) to return 50 to 60 visual sensitivities regularly spaced across the central 30 degrees of an eye in either a rectangular or polar grid. While the spatial pattern of damage exposed by such tests are useful for diagnosing and monitoring eye disease, summary metrics like Mean Deviation (MD) ^1^ or Mean Defect ^2^ are often used as a convenient summary of the field. These summary metrics are computed as a simple arithmetic average of the deviation of measured sensitivities from a normal population mean (total deviation, TD) and sometimes weighted by population variance as in the case of MD^3^. The simple average is a meaningful representation of the underlying true field as the locations at which sensitivity is measured are evenly spaced; thus, each location samples the same amount of spatial area in the field. If we test on irregular grids, this is no-longer the case, and so weighting locations by their spatial contribution to the average is more meaningful. We demonstrate this with an example in the next section.

While useful for cross sectional diagnosis and staging of disease, one of the major uses of summary measures are for monitoring change over time. This is often achieved by looking at the slope of linear regression through summary metrics of a series of fields over time to get a rate of change in dB/year ^1,4^. This approach works well on fixed grids as every location at a visit has a measurement from the previous visit (except the first visit) thus looking at the simple difference from visit to visit is meaningful. If the test pattern changes between visits, it is not necessarily the case that a simple difference between visits will represent a change in vision; it could be biased by the addition or removal of test locations. Examples of situations where this might occur is swapping to and from the 24-2 to 24-2C pattern ^5^, or using methods designed to adapt visual field test patterns based on prior results of an individual to delineate scotoma borders with higher fidelity ^6–8^. We demonstrate this with an example in the next section and propose a solution. Finally, we apply the new method to some artificial fields to demonstrate the effectiveness of the approach.

## Methods

### Motivating example

Consider the simple motivating example of an eye that is known to be unchanging over 6 visits with most of the field having locations with a TD of 0 dB except for a superior arcuate defect where all TDs are −30 dB. Figure 1A shows the visual field using a grid with 2 degree spacing covering the common rectangular 24-2 grid pattern (Humphrey Field Analyzer, Zeiss). Note that for this motivating example we are assuming that the field is perfectly measured at each visit; the introduction of measurement noise makes no substantial differences to the methods discussed, although the exact numbers may change if noise is added. Beginning with the 24-2 pattern at visit 1, 10 locations are added to the test pattern in the superior hemifield at each visit. Figure 1B and 1C shows the tested locations at visits 1 and 6.

**Figure 1.**
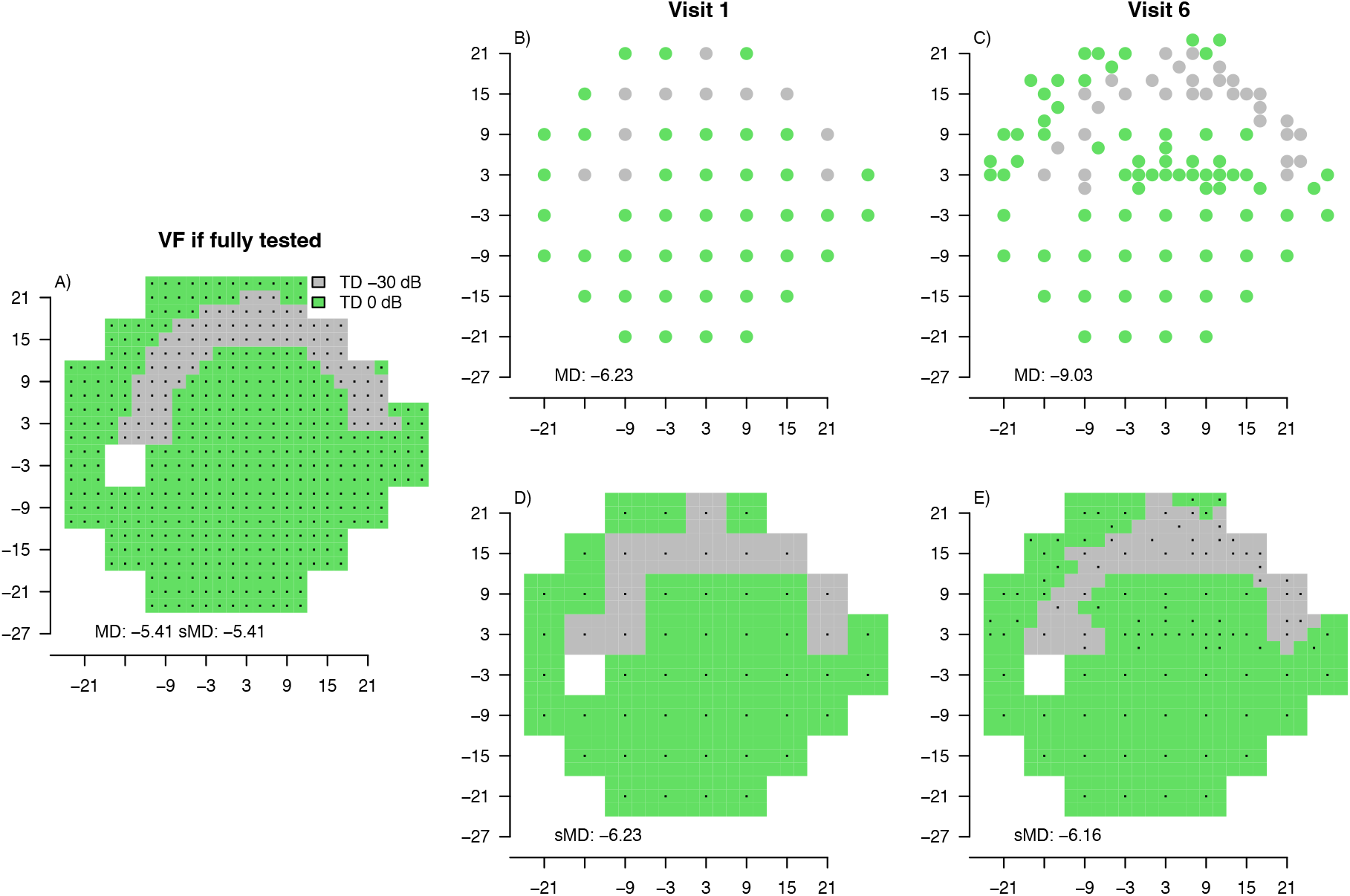
Motivating example of a stable eye with an arcuate defect as shown in A at all visits. Green represents a TD of 0 dB and grey TD of −30dB. B) The 24-2 pattern measured at the first visit (53 locations). C) The test pattern measured at visit C (103 locations). Panels D and E show the spatial area covered by test locations. MD is the average of TD values measured. sMD is the spatially weighted average of the TD values measured.

If we compute the simple average of deviations at each visit (MD), we get a value of −6.23 dB for Visit 1 and −9.03 dB at Visit 6. Although the underlying true visual field is not changing at all at each visit, the mere act of measuring more locations and including them in the average with equal weight leads to a decrease in the global average.

As mentioned in the introduction, this is mitigated by spatially weighting each location in its contribution to the average. We denote the spatially weighted average of TD values as sMD. If we use the number of closest 2×2 squares to weight each measured location, as shown in panels D and E of Figure 1, then the global averages become −6.23 dB and −6.16 dB at visits 1and 6 respectively. The known MD and sMD of the field as shown in panel A is −5.41dB, so as expected the more test locations that are added, the more likely sMD approaches the true underlying value.

If we were to compute linear regression on both MD and sMD for this eye over visits 1 to 6, assuming equally spaced visits as the x-axis, we get rates of change of −0.48 dB/visit (p = 0.001) and+0.03 dB/visit (p = 0.891). Neither of these are convincingly 0 dB/visit, the true rate of change.

### Linear rates of change

One of the reasons that regression on sMD in the motivating example does not accurately reflect the true rate of change is that the first time a location is measured it can contribute to a change in sMD that has not been measured. Consider a location that is introduced to the test pattern at visit *i* > 1; it contributes a change in sMD proportional to the difference between its implicitly assumed value at visit *i*− 1 and its measured value in visit *i*. Note the change itself has not been measured but still contributes to sMD. In the motivating example, one such location is (7, 21), which has an assumed TD value of 0 dB for visits 1 to 4, but when the location is measured at visit 5 it is −30 dB, contributing towards a lowering of sMD, hence a lowering of the regression slope computed on sMD.

One way of avoiding this potential distortion of change in sMD is to only include locations that have been previously measured in the computation of the slope on sMD. This is not as simple as excluding these values from the computation of sMD at each visit because while a value first tested at visit *i* is not needed in sMD for the computation of change from visit *i*− 1 to *i*, it is needed for the computation of change from visit *i* to *i* + 1 and also for reporting the measured sMD at visit *i*.

Thus, at each visit we must compute two aggregating values: sMD, the spatially weighted average of all locations as above; and a second, denoted sMD’, which is the spatially weighted average excluding any locations that were not tested in the previous visit. Linear regression slope is then computed from the slope between sMD’ at visit *i* and sMD at visit *i*− 1 for all visits *i*. This calculation is facilitated by rewriting the usual slope calculation as follows.

Assuming we have *n* visual field measurements *y*_*i*_ collected at time *x*_*i*_, 1 ≤ *i* ≤ *n*, the slope of linear regression is often defined as

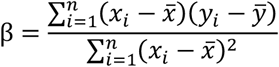

This can be rewritten using Abels Lemma ^9^ also known as Summation by Parts ^10^ as a weighted sum of the slope between each time point as

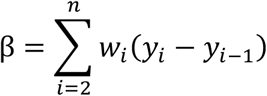

where

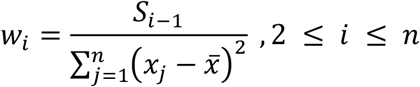

and

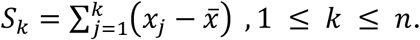

In our case, we use *y*_*i*_ taken from sMD’ and *y*_*i*−1_ taken from sMD at each visit *i* as shown by the dashed lines in Figure 2.

**Figure 2.**
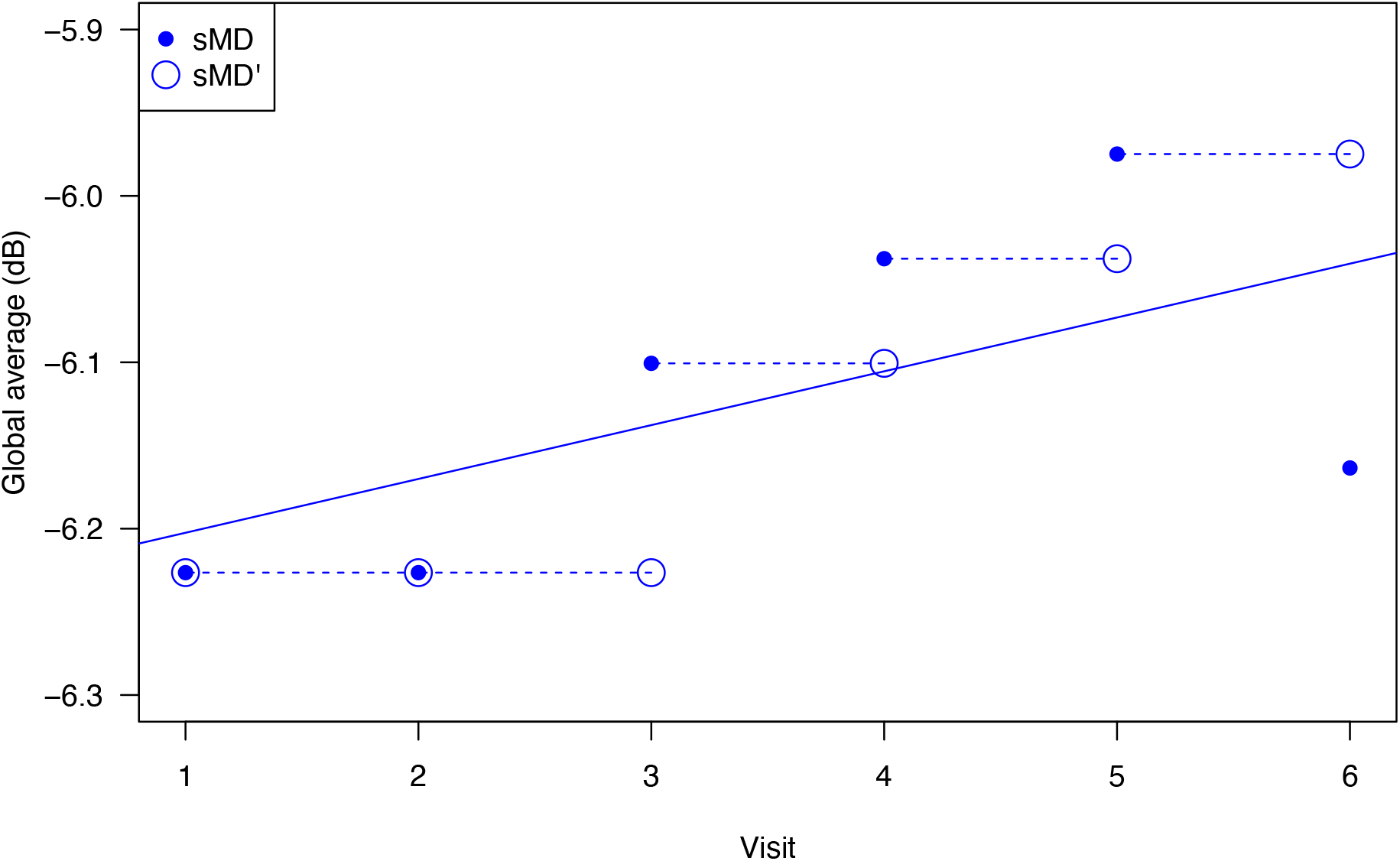
Linear regression at Visit 6 of the motivating example in Figure 1 (a stable, non-progressing eye). Solid blue symbols show sMD and the solid blue line linear regression on sMD. Dashed lines show the y_i_ − y_i−1_used for computing slope based on sMD+sMD’.

Table 1 shows the regression slopes computed on MD, sMD alone, and the combination of sMD+sMD’ which excludes locations that were not present in the previous visit. As can be seen, using sMD+sMD’ gets the correct slope of 0 db/visit in this example and is a perfect fit for the data, hence the p-values produced using a one-sided permutation test in the final column of the table are undefined.

**Table 1.** Global metrics computed for the example in Figure 1. MD – simple average; sMD spatially weighted average. Slopes are computed using linear regression with the p-value being from a one-sided test on slope < 0. The final column is a perfect fit (all residuals are 0), so the p-value is undefined.

| Visit | MD (dB) | sMD (dB) | Slope MD (dB/visit) | p | Slope sMD (db/visit) | p | Slope using sMD + sMD'(dB/visit) | p |
| --- | --- | --- | --- | --- | --- | --- | --- | --- |
| 1 | -6.23 | -6.23 |  |  |  |  |  |  |
| 2 | -7.14 | -6.23 | -0.92 | - | -0.00 | - | 0.00 | - |
| 3 | -7.81 | -6.10 | -0.79 | 0.029 | 0.06 | 0.833 | 0.00 | - |
| 4 | -7.95 | -6.04 | -0.58 | 0.021 | 0.07 | 0.973 | 0.00 | - |
| 5 | -8.06 | -5.97 | -0.45 | 0.012 | 0.07 | 0.997 | 0.00 | - |
| 6 | -9.03 | -6.16 | -0.48 | 0.001 | 0.03 | 0.891 | 0.00 | - |

### Experiments

To examine the general applicability of the method for calculating rate of change, and what the incorporation of spatial weighting might mean in practice, we simulated series of visual fields over 6 visits and compared the slope of regression on MD, sMD and sMD+sMD’ for each series against the underlying true value. The universe of possible locations for all fields were the 477 locations defined by a rectangular grid with 2 degree spacing inside the convex hull of the 24-2 pattern excluding a 6×6 degree rectangle located at (−15, 3) for the blind spot (the area in Figure 1A). All locations in the universe were uniformly randomly assigned a “true” starting value between −1 dB and 35 dB inclusive. We simulated 3 field types.

1. Stable fields where all locations kept the same value for all 6 visits.
2. Progressing fields where a uniform random number of locations in the range 3 to 45 were changed at each visit by a fixed, uniform random amount in the range −4 dB to +0.1 dB/visit. When a location is selected for change at a visit, that rate of change applies for that location in all future visits. Locations that progressed below −1 dB were set to −1 dB at any visit. For each field type we generated 50 series and then “tested” each series 50 times. Each test assumed no noise in the visual field measurement and added a uniform random number of locations in the range 3 to 10 per visit to the test pattern. Once a location was added to the test pattern, it remained in the test patter for all remaining visits. Total deviations were simply calculated as the location’s measured value less 30 dB.

## Results

Figure 3 shows the error in slope estimation from the ground truth on the 50 simulated progressing eyes each measured 50 times at visits 3, 4, 5 and 6 with both the location of damage and testing pattern uniformly randomly selected. The grey box in the right of each panel shows the error in slope of MD collected using the 24-2 pattern. The remaining 3 boxes in each panel show the error in slope computed on the test patterns with increasing number of locations. The white box is a simple average of all tested points at each visit and illustrates why simple averaging is not appropriate. As expected, the simple average has a wide range of errors in slope estimation because included in the average are location that have first been tested within the series and locations are not spatially weighted. The red box adds spatial weighting to the computation of the average to get sMD but still uses standard linear regression which counts new locations as a change contributing to the slope. Similarly, this does not work well. Finally, the blue box uses the recommended method presented in this paper, spatially weighting locations and excluding new locations from the regression at their first appearance (sMD+sMD’). As can be seen, the errors in estimating global change are in the same range as if a fixed 24-2 pattern was used. Using a paired t.test between grey and blue at each visit revealed a small (all < 0.003 db/visit) decrease in mean error for slopes estimated using variable grids at each visit (all p < 0.0001). The results for the stable eyes follow a similar pattern, but with almost all errors being 0 for both the sMD+sMD’ and 24-2 MD methods (not shown).

**Figure 3.**
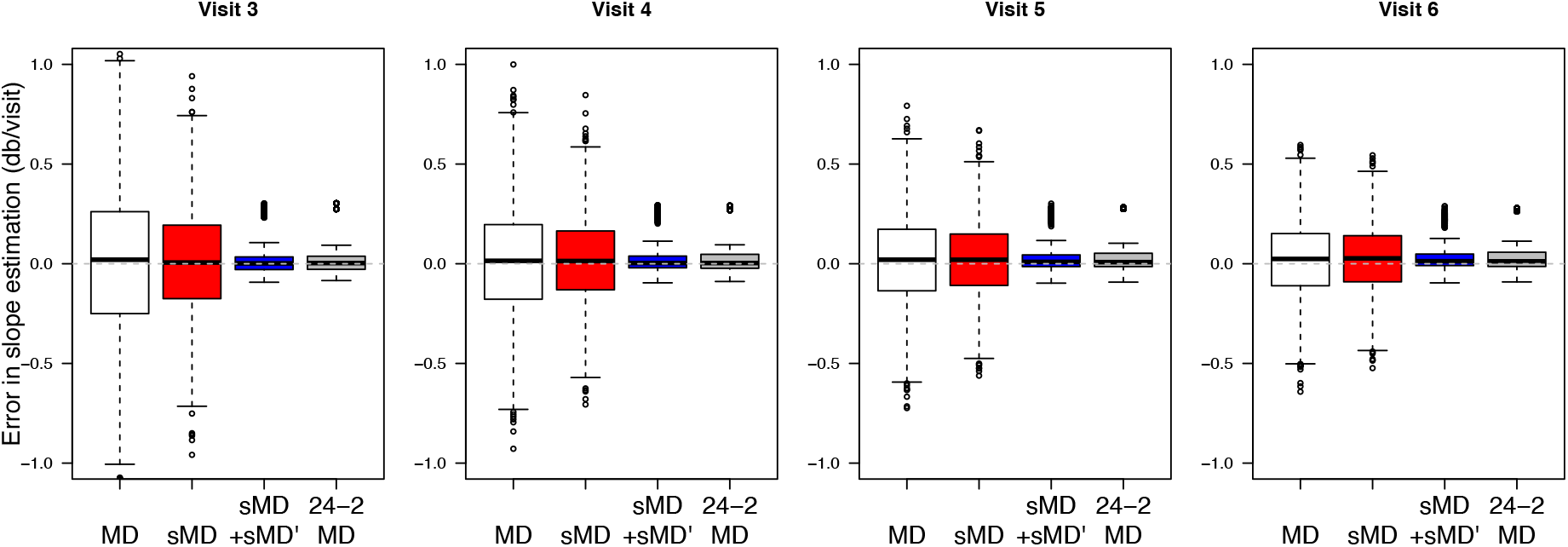
Error in slope estimation for progressing eyes at visits 3 to C pooled over all eyes and repeated measurements using different methods for computing averages as input to the regression. The test pattern is fixed at 52 locations for the 24-2 method; all others have between 3 and 10 locations added at each visit (see Methods). The MD uses a simple average of TD values as input to the regression; sMD uses spatial weighting of the average; and sMD+sMD’ is the method discussed in the text for ignoring differences between newly tested locations and the previous visit in the regression. Boxes show 25^th^ and 75^th^ percentiles, with whiskers extreme values up to 1.5 times the box length (default R boxplot).

## Discussion

In this paper we have introduced a method for calculating linear rates of change on spatial averages for series of visual fields that have differing test patterns from visit to visit. While the focus has been on “global” averages that use all tested locations at each visit, the methods described will work on spatial averages at any scale, for example, cluster averages.

We showed that when run on a visual field series with uniformly selected random locations progressing and test patterns evolving at each visit by adding uniformly selected random locations, the method offers estimates of the rate of change with errors the same as for a fixed 24-2 pattern (Figure 3). This is to be expected as with evenly spread change a regular pattern of test locations is the best one can do. This is one of the reasons that fixed patterns like the 24-2 have been used since the birth of automated perimetry; for general diagnosis without any prior information of possible damage, a regularly spaced grid is optimal for testing. It is comforting, therefore, that the new method produces similar rates of change as for regression on the average of the 24-2 pattern at each test.

Unlike the random examples used in our tests, in practice it is likely that progressing locations will have some spatial pattern and that test patterns will change over visits using some intelligent logic. In this more realistic scenario, we would expect the evolving test patterns with rates calculated using sMD+sMD’ to be a more realistic representation of the true underlying rate of change of the whole field.

As an aside, we note that the discourse above focusses on adding test locations to a test pattern over time, but the sMD+sMD’ method for estimating linear rates of change works equally well when points are removed from a test pattern, or when there is a mix of removing or adding test locations.

## Data Availability

All data produced in the present work are contained in the manuscript

